# Cost-effectiveness and return on investment of protecting health workers in low- and middle-income countries during the COVID-19 pandemic

**DOI:** 10.1101/2020.06.22.20137257

**Authors:** Nicholas Risko, Kalin Werner, O. Agatha Offorjebe, Andres I. Vecino-Ortiz, Lee A. Wallis, Junaid Razzak

**Author notes:** Corresponding Author: Dr. Nicholas Risko, MD. Author Contributions: NR, KW, and AVO contributed to model development. NR, KW, and OAO contributed to model inputs and outputs. NR, KW, AVO, LW and JR contributed to data interpretation, framing and writing. Funding: No funding to disclose.

## Abstract

**Background:** In this paper, we predict the health and economic consequences of immediate investment in personal protective equipment (PPE) for health care workers (HCWs) in low- and middle-income countries (LMICs).

**Methods:** To account for health consequences, we estimated mortality for health care workers (HCW), and present a cost-effectiveness and return on investment (ROI) analysis using a decision-analytic model with Bayesian multivariate sensitivity analysis and Monte Carlo simulation. Inputs were used from the World Health Organization Essential Supplies Forecasting Tool and the Imperial College of London epidemiologic model.

**Results:** An investment of $9.6 billion USD would adequately protect HCWs in all LMICs. This intervention saves 2,299,543 lives across LMICs, costing $59 USD per HCW case averted and $4,309 USD per HCW life saved. The societal ROI is $755.3 billion USD, the equivalent of a 7,932% return. Regional and national estimates are also presented. In scenarios where PPE remains scarce, 70-100% of HCWs will get infected, irrespective of nationwide social distancing policies. Maintaining HCW infection rates below 10% and mortality below 1% requires inclusion of a PPE scale-up strategy as part of the pandemic response.

**Discussion:** In conclusion, wide-scale procurement and distribution of PPE for LMICs is an essential strategy to prevent widespread HCW morbidity and mortality. It is cost-effective and yields a large downstream return on investment.

## Background

On March 3, 2020, eight days before the World Health Organization (WHO) declared coronavirus 2019 (COVID-19) a global pandemic, there was already concern about depleted global stock of personal protective equipment (PPE). At that time, the WHO estimated a need to increase worldwide production by 40% to provide monthly requirements of 80 million masks, 76 million gloves, 30 million gowns, and 1.59 million goggles [1]. As COVID-19 swept parts of East Asia, Europe and the United States (US), it became evident that even in resource-rich health systems there was inadequate supply of PPE to protect frontline health care workers (HCWs)[2]. A survey conducted by the US Association for Professionals in Infection Control and Prevention in late March found that 48% of healthcare facilities were out or nearly out of N-95 respirators and only 32% reported having sufficient gowns [3]. The supply shortages have led to PPE rationing and reuse of equipment beyond manufacturer recommendations [4]. The US Centers for Disease Control and Prevention (US CDC) and the WHO have released recommendations for optimizing PPE supply [5,6] and the US Food and Drug Administration has taken the unprecedented step of issuing emergency approval for sterilization techniques to allow reuse of previously disposable PPE [7]. The resulting global bidding war, exportation restrictions and supply chain disruptions have hit low- and middle-income countries the hardest [8,9].

Over 80% of the world’s population lives in LMICs where fragile health systems with few resources make HCWs vulnerable to COVID-19 [10,11]. Given the pre-existing shortage of HCWs [12], even minimal workforce depletion due to illness, death or absenteeism could threaten the stability of LMIC health systems. The COVID-19 pandemic has yet to reach its peak in many of these countries, however they are already bracing for the potential of a long and devastating disaster. This paper presents the findings of a cost-effectiveness and a return on investment analysis to determine the health and economic impact of immediate scale-up in the production and distribution of PPE for 139 LMICs. The aim is to inform active global, regional and national discussions on policy, strategy and financing to protect HCWs and the integrity of health systems in LMICs.

## Methods

We developed a decision-analytic model to compare the costs and effects of two PPE use scenarios at the global and regional levels for all LMICs, following standard guidelines for cost-effectiveness analyses [13,14]. A base case where full PPE supply maintains a low rate of HCW infection was compared to a scenario where inadequate PPE leads to higher rates of HCW infection. Our main outcomes were: 1) cost per HCW death averted and 2) cost per HCW case averted. A Return on Investment (ROI) analysis was also performed by comparing the societal economic gains from having HCWs fully protected against exposure with the current investment required to afford the PPE. Finally, we assessed the impact of the projected HCW infections and deaths on the estimated worker pool in each region.

To estimate PPE resource use and costs we utilized the WHO COVID-19 Essential Supplies Forecasting Tool (ESFT) [15]. The ESFT is designed to help governments and other stakeholders estimate essential supply requirements to respond to the COVID-19 pandemic. We ran projections for each LMIC for a 30-week period starting in mid-June and incorporated costs related to “hygiene” and “PPE” into our decision-analytic model. Ideal PPE availability is consistent with WHO best practice guidelines [6]. This implies gloves, gown, face shield and masks for all encounters involving a suspected case and enhanced precautions for aerosol generating procedures. PPE costs in EFST are intended to directly inform procurement and reflect competitive market prices in the global market. The EFST does not provide confidence intervals for their costing data, so we created sampling bounds for our sensitivity analysis informed by variations in bulk order market prices.

The costs of labor and healthcare utilization were taken from WHO-CHOICE [16]. Costs were tabulated in 2020 US dollars (USD) from the societal perspective. Consistent with this, lost future productivity due to early mortality was included in assessment of the economic impact. Training costs, whether viewed as a lost investment in HCWs that have died or as a replacement cost to replenish the work pool, were not included due to the difficulty of estimating this for each setting globally. Their absence has likely led us to underestimate the economic benefit of averted mortality. Our tool and the EFST align with standard international definitions of who constitutes a HCW [12,15]. In general, HCWs are professional members of the healthcare community that will interact closely with a patient, such as doctors, nurses, technicians/medics, and ancillary staff.

The ESFT utilizes a basic Susceptible-Infectious-Removed (SIR) model that is described in the tool [15]. The tool was run on default settings using incorporated background data for each country, the tally of current cases in each country as of June 6, a medium clinical attack rate of 20%, a targeted testing strategy for all severe/ critical patients, and 10% of mild/moderate cases being tested. In addition, we incorporated estimates of national mortality and hospitalizations from published projections calculated by the WHO Collaborating Center for Infectious Disease Modeling at the Imperial College of London (ICL) [17]. The three scenarios presented in the ICL model, including unmitigated pandemic spread, suppression with intensive social distancing after reaching a trigger of 1.6 deaths per 100,000 population per week, and suppression after reaching 0.2 deaths per 100,000 population per week were analyzed for their varying impact on case and mortality counts. This informed our ranges for Bayesian sensitivity analysis and our exploration of policy impact on workforce depletion.

PPE use decreases transmission of aerosolized respiratory viruses and PPE shortages will be associated with elevated HCW infection rates, however uncertainty remains around the exact level of impact [18,19]. Due to the inherent difficulty of directly measuring the real-world efficacy of PPE, we used the variation in the proportion of HCW infections to total infections, as a proxy indicator for the quality of protection in the workplace. Available figures demonstrate a range from nearly 0% to over 20% [20-30]. Given this high level of uncertainty, for the comparator case with low-PPE availability we designed the model to randomly sample from a wide range of infection rates (4.5-25%) during the 10,000 run Monte Carlo simulation. **Table 1** presents key parameter values, their ranges of uncertainty, their distribution for Bayesian analysis and their sources.

**Table 1:**
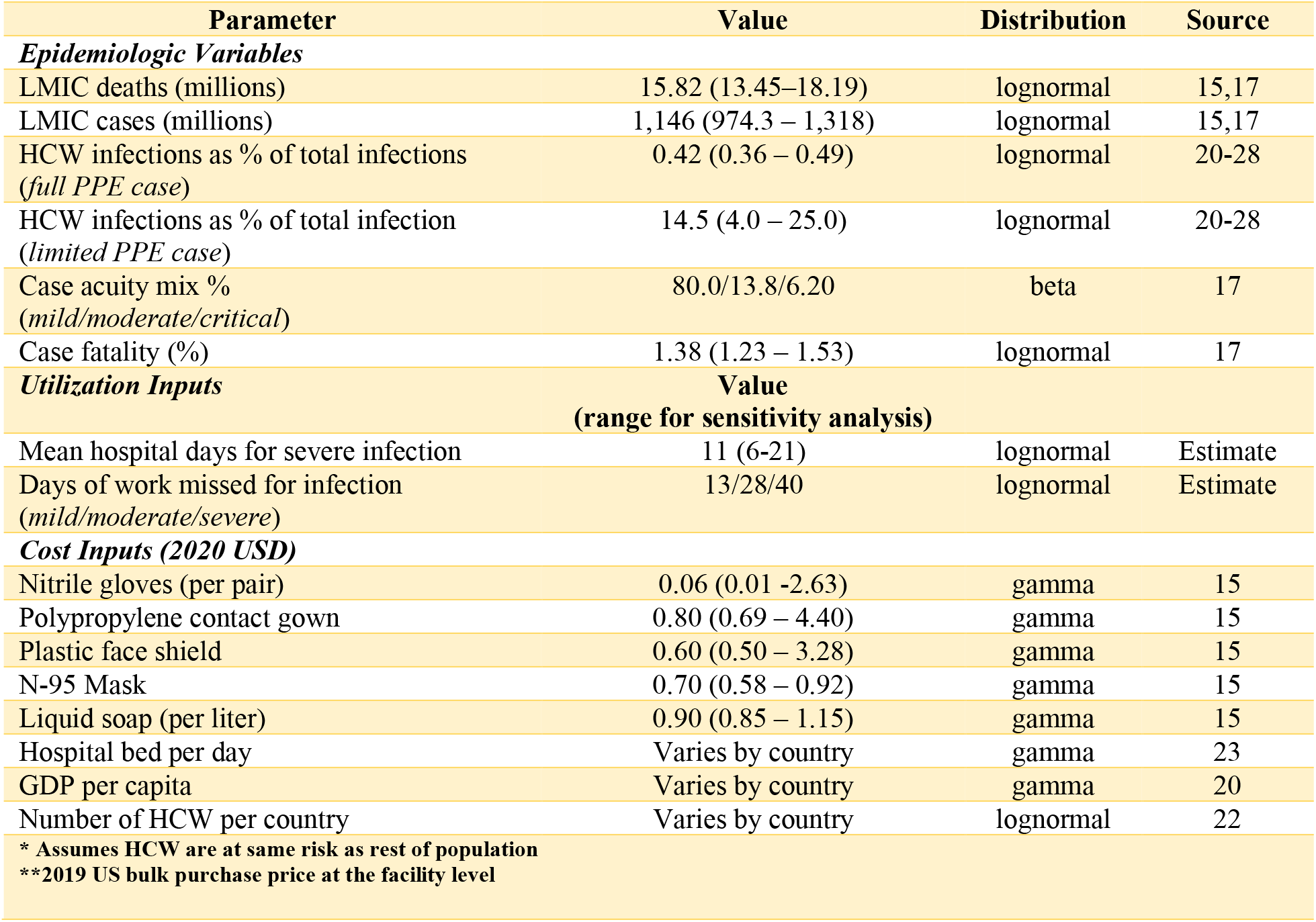
Key model parameters.

### Sensitivity Analysis

We performed a Bayesian multivariate sensitivity analysis to consider the uncertainty surrounding all key parameters. A 10,000 run Monte Carlo simulation randomly re-sampled across the input distributions for each model parameter for each regional projection. Beta distributions were used for sampling within the 95% confidence interval of probability variables, gamma distributions for cost variables and lognormal distribution for the remaining parameters. **Figure 1** contains cost-effectiveness plane scatter plots for both the number of cases averted, averted mortality for all LMICs, along with breakdown by World Bank Region for 139 LMICs in East Asia and Pacific (EAP), Europe and Central Asia (ECA), Latin America and Caribbean (LAC), Middle East and North Africa (MENA), South Asia (SA) and sub-Saharan Africa (SSA). **Figure 2** presents the ROI curves for each region generated by the Monte Carlo simulation.

**Insert Figure 1:**
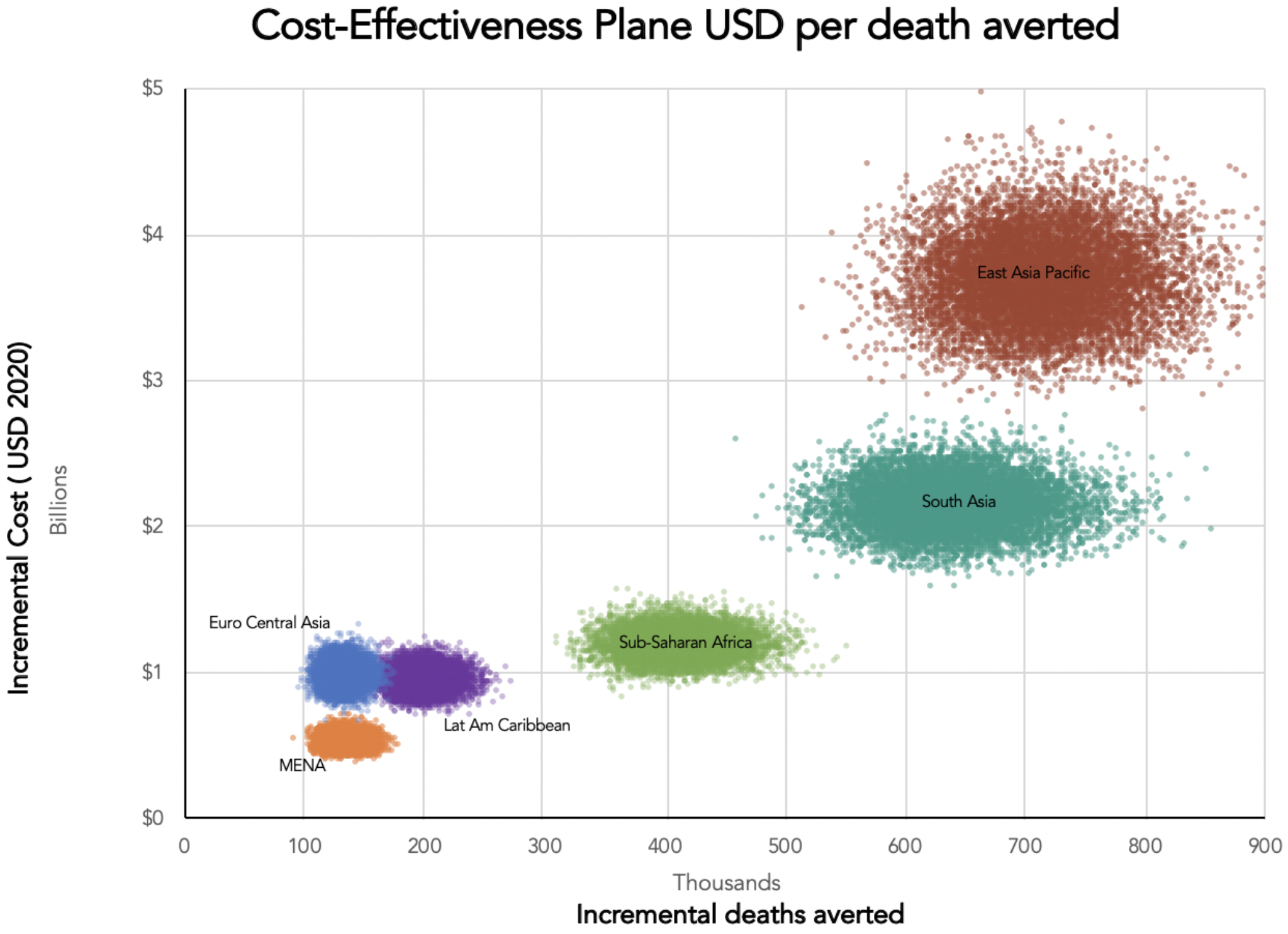
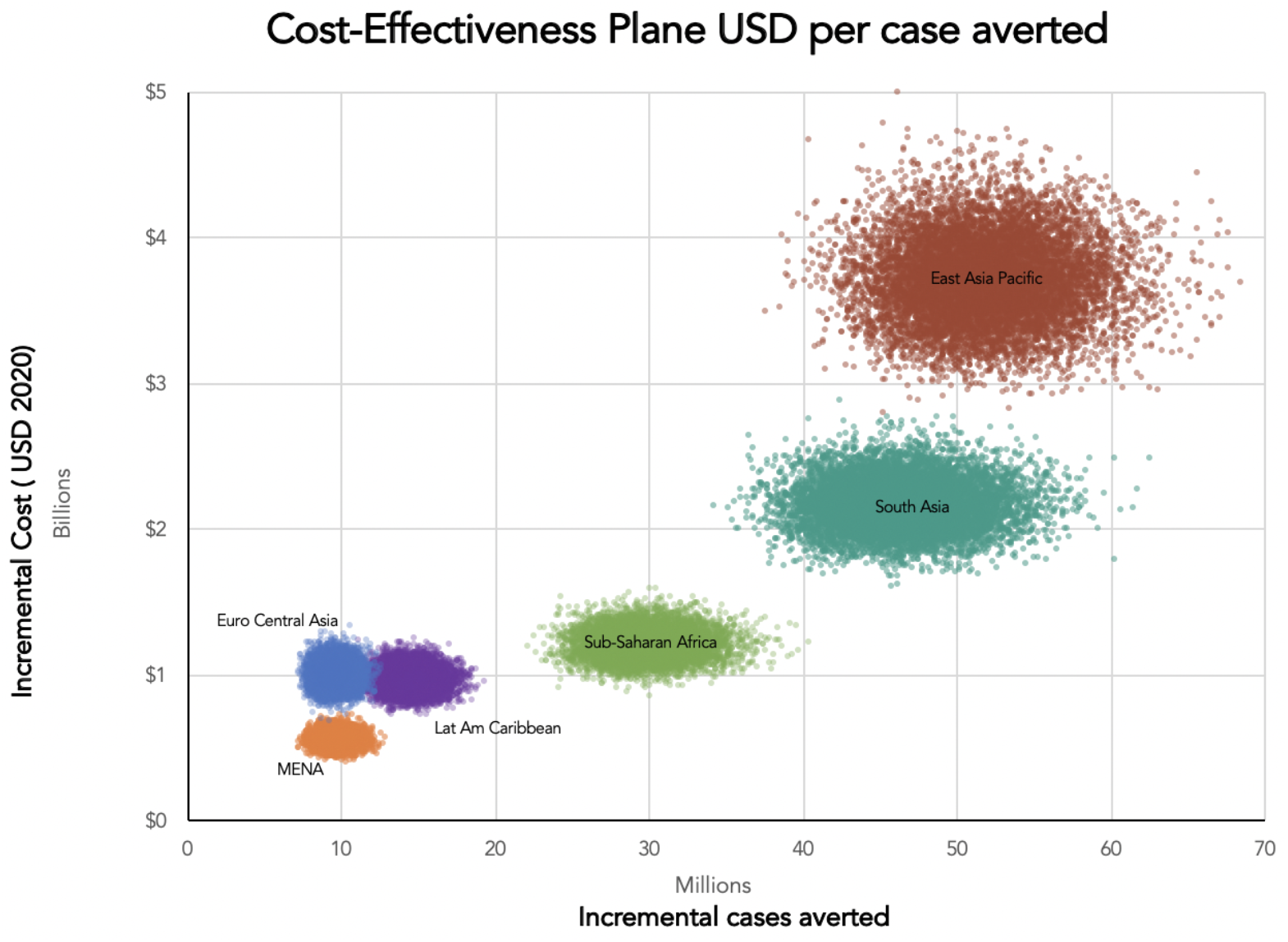
Cost-effectiveness planes by region.

**Insert Figure 2:**
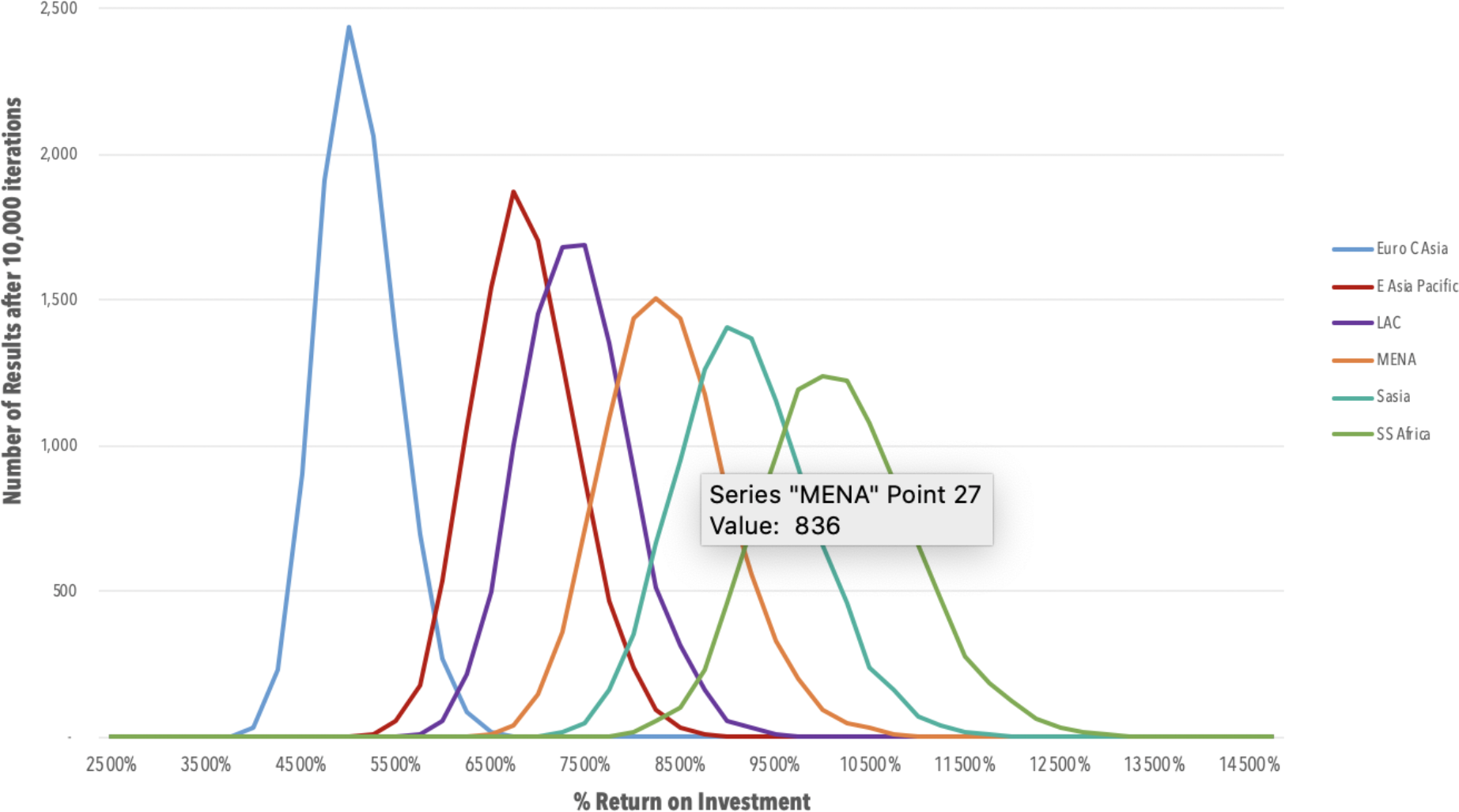
Return on investment curves by region.

## Results

The model predicts that across all LMICs there will be 166,689,862 HCW cases and 2,299,543 deaths if PPE supply remains constrained. Purchasing and distribution of PPE to allow for adequate protection of all HCWs requires an investment of $9.6 billion USD. This would result in a reduction to 4,863,299 HCW cases and 67,283 HCW deaths, saving roughly 2,232,260 lives with a mean incremental cost-effectiveness ratio of $59 USD per HCW case averted and $4,309 USD per HCW life saved. The societal ROI from productivity gains is estimated to be $755.3 billion USD, yielding the equivalent of a 7,932 % ROI. Breakdown by World Bank Region can be found in **Table 2**.

**Table 2:** Summary of cost-effectiveness results*.

| <u>Incremental Change</u> |  |  |  | <u>Cost-effectiveness Ratios</u> |  |  |
| --- | --- | --- | --- | --- | --- | --- |
| Region | HCW Cases Averted<br>(in millions) | HCW Deaths Averted | Investment<br>(in millions) | Cost per Case Averted | Cost Per Death Averted | Economic Gains<br>(in millions) |
| East Asia & Pacific | 51.9<br>(49.3 to 54.5) | 713,277<br>(677,963 to 748,590) | \$3,711<br>(3,526 to 3,895) | \$72 (67 to 78) | \$5,237<br>(4,862 to 5,611) | \$257,421<br>(247,433 to 267,407) |
| Europe & Central Asia | 9.61<br>(9.11 to 10.1) | 132,632<br>(125,831 to 139,433) | \$993.4<br>(946.2 to 1,040) | \$104 (97 to 111) | \$7,541<br>(7,014 to 8,069) | \$51,769<br>(49,839 to 53,698) |
| Latin America & Caribbean | 14.5<br>(13.7 to 15.2) | 200,069<br>(189,920 to 210,219) | \$959.9<br>(914.6 to 1,005) | \$67 (62 to 71) | \$4,830<br>(4,496 to 5,164) | \$72,125<br>(69,623 to 74,986) |
| Middle East & North Africa | 9.72<br>(9.25 to 10.2) | 133,895<br>(127,364 to 140,427) | \$544.7<br>(518.7 to 570.6) | \$56 (53 to 60) | \$4,094<br>(3,811 to 4,376) | \$46,024<br>(44,187 to 47,865) |
| South Asia | 46.4<br>(44.1 to 48.7) | 640,080<br>(608,652 to 671,507) | \$2,158<br>(2,056 to 2,260) | \$47 (44 to 50) | \$3,393<br>(3,163 to 3,623) | \$200,343<br>(191,551 to 209,135) |
| Sub-Saharan Africa | 29.8<br>(28.4 to 31.3) | 412,148<br>(392,387 to 431,909) | \$1,202<br>(1,144 to 1,259) | \$41 (38 to 43) | \$2,934<br>(2,735 to 3,132) | \$123,442<br>(117,922 to 128,961) |
| LMIC aggregated | 161.8<br>(153.9 to 169.8) | 2,232,260<br>(2,122,083 to 2,342,436) | \$9,557<br>(9,100 to 10,014) | \$59 (55 to 63) | \$4,309<br>(4,010 to 4,608) | \$755,314<br>(724,335 to 786,293) |

95% confidence intervals are derived using the standard error of the simulation results
\*All monetary values are in 2020 US dollars, rounded to nearest dollar.

**Figure 3** illustrates the estimated percentage of the HCW pool infected under various scenarios. **Figure 4** examines mortality as a percentage of the HCW pool. The unmitigated, mitigated (1.6 deaths/100,000 population/week trigger) and suppressed (0.2 deaths/100,000 population/week trigger) labels correspond to the three levels of social distancing in the ICL model. In scenarios where PPE remains scarce and there is less than full suppression, 100% of HCWs are projected to get infected. Keeping the proportion infected under 70% requires incorporation of PPE scale-up into overall public health strategy. Likewise, the inclusion of a PPE strategy significantly reduces mortality.

**Insert Figure 3:**
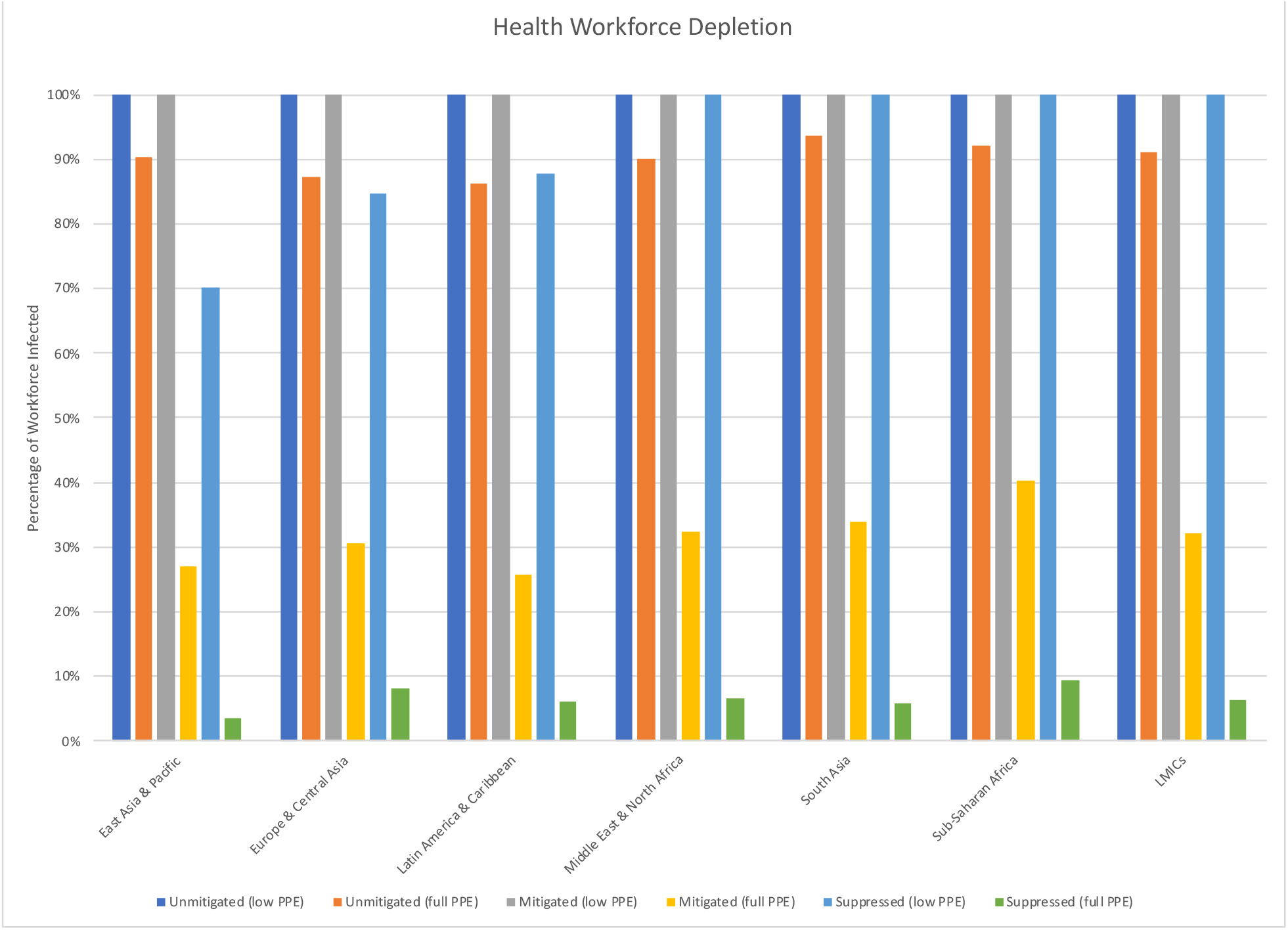
Cumulative HCW cases as a percentage of total workforce, by strategy.

**Insert Figure 4:**
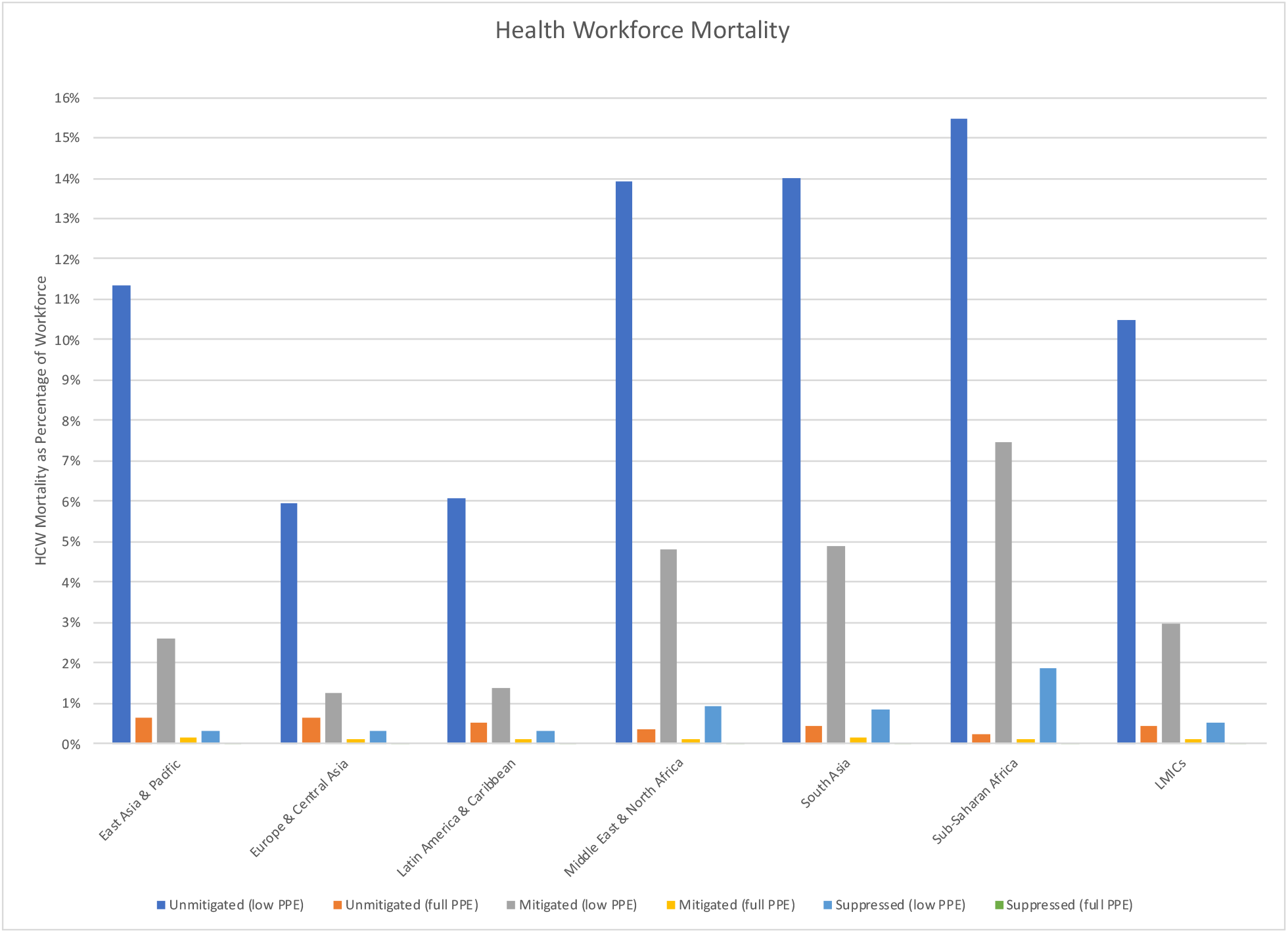
Cumulative HCW mortality as a percentage of total workforce, by strategy.

## Discussion

The global shortage of PPE has become a critical issue, given the high risk of COVID-19 transmission to HCWs during care encounters. Scarcity is highest in LMICs, where projections estimate the pandemic’s impact will be the heaviest. Utilizing data regarding variations in HCW infection rates across PPE availability scenarios alongside the WHO ESFT, we projected the cost-effectiveness and ROI of PPE scale-up.

Our PPE demand forecasts, costs, infection and mortality estimates are driven by projections from ICL and the EFST, whose methods and limitations have been previously described [15,17]. Given the speed of this pandemic, further limitations exist in the availability of data examining the effect of PPE on HCW infection rates, as well as the true percentage of total cases that are HCWs. To inform our model we relied on a collection of sources that include rigorous scientific studies, data published by government entities, and media reports that cite government sources. Overall, data are consistent in showing low to zero HCW infection rates in highly controlled settings with strict PPE compliance while public health figures that incorporate a range of healthcare settings with varied PPE compliance demonstrate rates as high, or above 20%. Examples of the percentage of cases that are HCWs from a variety of settings include: 0-1% in high PPE compliance scenarios in Hong Kong, China and the Netherlands [20-22]; 19.9% in the United States [23]; 12.2% in Italy [24]; 3% in South Africa [25]; 7.5% in Nigeria [26]; 13% in Egypt [27]; and 18% in the Philippines [28]. In another Chinese example, initial studies estimated that up to 41% of cases were acquired in the healthcare setting; however, later reports after enforcement of stringent PPE guidelines show HCW infections comprised 3.8% of the total cases [29]. One organization estimates that globally, HCWs make up 7% of all cases [30].

In the absence of perfect data, we have endeavored to make all assumptions as conservative as possible and to rigorously explore them in our sensitivity analysis. Whenever possible, our approach was to allow this real-world uncertainty to exist in the model. For example, our comparator case uses a wide range of HCW infection rates (from 4.5-25%), from which the model samples during the Monte Carlo simulation. Furthermore, we have not integrated the downstream effect on future patients of keeping HCWs safe and healthy into our cost-effectiveness analysis. In locations where workforce depletion could affect the quality of healthcare service, there is likely to be a substantial positive externality extending beyond the individual HCW.

Unfortunately, fierce competition in the global PPE market has led wealthy countries to outbid poorer ones and, in some cases, activate legislation preventing exportation of domestically produced PPE.[31] Global financing mechanisms have been developed to provide COVID-19 relief to LMICs; however, it remains unclear if funding will be earmarked for PPE and if resources will be adequate to protect HCWs.[32] It is likely that national governments will need to take proactive measures towards procuring and producing PPE. In a time of global economic recession, it appears this is an investment that can yield significant returns.

There are strong reasons for societies to protect their HCWs. Our analysis addresses only one of these reasons: immediate investment in the wide-scale production and distribution of PPE for LMICs yields a significant benefit in lives saved and ROI. Our findings also suggest it is a required component of public health strategy in order to prevent massive depletion of the workforce.

## Data Availability

Input data and cost-effective modeling is available upon request to the corresponding author

